# The epidemiology of Brucellosis and Q fever in a cross-sectional serosurvey of occupationally exposed groups in peri-urban Lomé, Togo

**DOI:** 10.1101/2024.10.28.24316261

**Authors:** Charlotte L. Kerr, Akouda Patassi, Pidemnéwé S. Pato, Javier Guitian, Sylvie Audrey Diop, Punam Mangtani, Patrick Nguipdop-Djomo

**Author notes:** Author Contributions: CK and PND conceived and designed study. PM, SAD, PP and AP contributed to the design. CK, JG and PP developed the sampling frame and recruitment. CK performed data collection and analysed data with supervision from PM and PND. AP and PP provided in-field support and resources. CK wrote a first draft of the paper. PND, PM, AP, SAD, JG and PP provided critical review of the paper.

## Abstract

**Background:** *Brucella* species and *Coxiella burnetii* have been detected in livestock populations in Togo. Populations exposed to livestock ruminants through occupation may be at increased risk of infection.

**Methods/Principal Findings:** A cross-sectional serosurvey was conducted in 108 abattoir and 81 dairy farm workers (from 52 dairy farms) in peri-urban Lomé, Togo in 2019-2020. Sera were tested using the Rose Bengal plate agglutination test (RBT) and the indirect Brucella IgG Enzyme-Linked Immunosorbent Assay (ELISA) for Brucella, and the IgG ELISA for *Coxiella burnetii* in Phase 1 and in Phase 2. Fresh bulk milk from farms were tested using an indirect milk ELISA for Brucella IgG.

Eighteen workers (9.5%, 95% CI 5.5-16.0) were Brucella seropositive. Twenty-eight percent (95% CI 22.5-34.3) of workers were seropositive for *C. burnetii*. Twenty of fifty-one farms which gave milk samples tested positive for Brucella (39.2%, 95% CI 26.6 - 53.4%).

Farmworkers had nearly twice the odds of being Brucella seropositive compared to abattoir workers (OR 1.93, 95% CI: 0.94-3.93, p=0.07). In farmworkers, working on farms with animal ill health, a positive milk test, participating in small animal husbandry and assisting with cattle abortion were all associated with increased odds of seropositivity. Workers who consumed unboiled milk at least every month were more likely to be seropositive (OR 3.79, 95% CI: 2.34-6.13, p<0.001) while participants who consumed fermented milk and cheese had greater odds of being seropositive for *C.burnetii* (OR 1.59, 95% CI: 1.26-2.00, p<0.001 and OR 1.70, 95% CI: 0.97-2.98, p=0.07 respectively).

**Conclusions:** Livestock workers in peri-urban Lome have been exposed to both *Brucella* and *Coxiella burnetiid* bacteria. The widespread consumption of unboiled dairy products and lack of PPE use is of concern as both dairy consumption and participation in animal husbandry activities have been seen to increase odds of seropositivity for both pathogens.

**Author summary:** Human and animal health are inextricably linked, particularly for those who live and work closely with animals. Brucellosis and Q fever are two zoonotic diseases transmitted through animal contact and dairy product consumption, which cause non-specific fevers and for which diagnostic tests are lacking in many LMIC contexts. Previous studies have shown that both bacteria circulate in livestock in Togo. We undertook a survey in dairy farm and abattoir workers in peri-urban Lomé, Togo, and found that 9.5% and 28% of workers were seropositive for *Brucella* and *C.burnetii* respectively. We found that risk factors included animal husbandry practices and consumption of dairy products. Mitigating practices such as the use of PPE and boiling milk are simple ways that livestock workers could protect themselves from these and other zoonotic disease.

## Introduction

Zoonotic pathogens maintained by ruminant reservoirs, such as *Brucella* species and *Coxiella burnetii*, are of concern in agricultural communities, particularly in low-income countries (LICs) where animals and humans frequently interact, infection status of livestock holdings is often unknown and controls are lacking. Workers in the livestock industry, including slaughterhouse workers, farmers of ruminants, animal healthcare workers and veterinarians, are particularly vulnerable to such zoonoses which not only impact health and wellbeing but also livelihoods through worker’s reduced capacity to do labour and through livestock productivity losses [1].

Both brucellosis and Q fever are neglected zoonoses associated with ruminant reservoirs, transmitted, to a varying extent, via contact with infected animals and their bodily fluids, consumption of animal products and inhalation of aerosols [2,3]. They present clinically as non-specific febrile illnesses which are often misdiagnosed as diseases such as malaria and typhoid fever due to lower awareness and lack of laboratory tests [2,4–7]. Both diseases can also result in more serious, chronic sequelae such as osteoarticular disease including spondylitis and osteomyelitis, neurological disease and endocarditis in Brucellosis, and hepatitis, pneumonia, heart disease and chronic fatigue in Q fever [5,8,9].

More than half of people in Togo work in agriculture, despite a rapidly urbanising population [10]. Local livestock production systems are largely informal and may magnify risk of exposure to these pathogens due to unrestricted grazing and transhumance, leading to a high-level of mixing between species and across large areas, and manual milking and slaughter of animals which is lacking hygiene measures [11].

A recent study looking at peri-urban dairy production in West and Central Africa found a Brucella herd prevalence of 62% in dairy herds from the surroundings of the capital city of Togo, Lomé [12]. *Brucella melitensis* was isolated from cattle hygroma samples in this region (personal communication). Another livestock study, in Northern Togo, found an individual *C.burnetii* seroprevalence of 16.1% in cattle, 16.2% in sheep, and 8.8% in goats [13]. However, there is a scarcity of good quality data on the level of human exposure to *Brucella* species and *C.burnetii* in Sub-Saharan Africa [7,14]. An increased focus on assessing the risk in these populations is required to control the burden of such preventable poverty-related diseases.

To our knowledge, no previous studies to ascertain the prevalence of *Brucella* or *C.burnetii* seropositivity have been undertaken in people who work with dairy cattle in Southern Togo. In this study we aim to focus on known at-risk populations, given the high prevalence of Brucellosis in large ruminants in the maritime region of Togo, to ascertain the burden of Brucellosis, and Q fever. We used a cross-sectional serosurvey in abattoir and farm workers in the peri-urban area of Lomé, Togo. We also assessed factors associated with higher odds of infection from these pathogens.

## Methods

### Study design and setting

The cross-sectional serosurvey in abattoir workers and livestock keepers was conducted between December 2019 and March 2020. It was embedded in a larger project on Brucellosis in animals in West and Central Africa which had surveyed 100 randomly sampled dairy farms in peri-urban Lomé in 2017 to 2018 [12]. Farmworkers were recruited from these and other farms in the three Western prefectures of the Maritime region which supply fresh cow milk to Lomé. Abattoir staff were additionally recruited at the municipal abattoir in Lomé.

### Participant selection

All staff employed by the abattoir and veterinary staff were selected for study participation given the small number. Of the 167 independent butchers registered at this abattoir and enumerated for this study 50% (84) were selected by simple random sampling with replacement when selected sampled workers were unavailable or refused.

Dairy farms from the larger project were preferentially enrolled, however due to herd movement, replacement farms for those who had left the area were selected by animal health workers using convenience sampling. Up to 3 individuals were randomly selected from a list of workers on each farm. An information leaflet describing the study was provided in French. A fieldworker explained in a local language where participants did not read French. Written consent was obtained prior to interview and blood collection. If the participant was illiterate a witness also signed the consent form. This sampling strategy was expected to be able to detect an estimated Brucella seroprevalence of 10% with 5% precision [13,15,16].

### Sample collection and processing

The phlebotomist collected 4mls of peripheral venous blood. Samples were labelled with unique identification numbers linked to questionnaire data, transported to the laboratory (Institut National d’Hygiene, Lomé) in a cool box at 4-8°C and centrifuged to obtain serum on the same day.

All sera were tested for anti-*Brucella* antibodies using the Rose Bengal plate agglutination test (RBT) (APHA, UK) and then stored in freezers at −20°C. This assay detects both agglutinating and non-agglutinating IgM, IgG and IgA antibodies [17]. All samples were also tested after sample collection was complete using the indirect Brucella IgG Enzyme-Linked Immunosorbent Assay (ELISA) (Serion ELISA classic, Germany) as per manufacturer’s instructions.

Sera were also analysed for detection of IgG antibodies against *Coxiella Burnetii* in either Phase 1, which predominates in chronic disease, or Phase 2, which predominates in acute disease, using the Serion ELISA classic (Institut Virion/Serion GmbH, Germany) and classified as positive, borderline or negative according to manufacturer’s instructions.

In addition, samples of fresh bulk milk were collected in 15ml plain tubes from each farm visited, transported in a cool box (4-8°C) to the laboratory (Department de l’Elevage, Lomé) where they were aliquoted into 1.5ml cryotubes and stored in freezers at −20°C. These were analysed as one batch using an indirect milk ELISA for *Brucella* IgG (APHA, UK).

### Data collection and study variables

In-person interviews were carried out in the participant’s language using a structured questionnaire with close-ended questions, on tablet computers using Open Data Kit (ODK)by trained interviewers.

The questionnaire consisted of three sections, respectively on farm-level risk factors, livestock contact, and consumption of livestock products.

Farm-level variables included number of animals on farm by species, morbidity and mortality in animals including abortion and hygroma, animal movements and mixing. Animal numbers were dichotomised into none or some, where ownership was rare, or using the median as cut-off.

Questions about livestock contact examined both husbandry and slaughter activity, with cattle, small ruminants and pigs examined separately, as well as asking about frequency of activity and duration, and use of protective equipment.

Consumption of liquid milk in any form, processed dairy products, and dried meat were noted, with products from cattle and from small ruminants assessed separately. Information on the frequency of consumption and whether products had been boiled or not was collected.

To deal with issues of scarcity for some variables a number of categories were merged. Assisting with cattle abortions was a rare occurrence and was classified as never or sometimes, whereas manual milking of cattle was commonly carried out on a daily basis and frequency was therefore classified as daily or less than daily/never.

Data were collected on potential confounders including age, sex, ethnicity, religion and education. Information was also gathered on recent health and healthcare-seeking behaviours, including recent episodes of pyrexia, muscle or joint pain and night sweats

The outcomes were positive *Brucella* or *C.burnetii* serology, defined as any individual who tested positive to either RBT or Brucella IgG ELISA for *Brucella*, and either Phase 1 or Phase 2 ELISA for *C.burnetii.* Borderline results were considered seronegative.

### Statistical analysis

Data were analysed using Stata 17 (Statcorp, College Station, TX, USA). Cross-tabulations were used to describe participants. Prevalence was calculated for both *Brucella* and *C.burnetii* seropositivity and 95% confidence intervals were computed using cluster-robust standard errors, with clustering at the site (individual farm/abattoir) level, and chi-square tests were used to compare clinical symptoms by serological status.

A hierarchical conceptual framework (Fig 1) was developed to guide the analysis, grouping potential risk factors from distal to proximal. Age and site of work (abattoir or farm) were considered *a priori* confounders. The association between seropositivity and farm-related exposure variables in farmworkers, slaughter-related exposure variables in abattoir workers, and animal products consumption in all participants, were respectively investigated using logistic regression, with robust standard errors to account for site-level clustering. The likelihood ratio test for heterogeneity was used to check for interaction between site of work (abattoir or farm) and the exposure variables. Due to limited statistical power in the data, multivariable analysis only examined the association between consumption variables and Coxiella seropositivity. The Wald test was used to assess evidence of association between the exposure variables and the outcome in the multivariable model. Models were assessed for multicollinearity and if detected the collinear variable which was least associated with the outcome was removed. Confounding was assessed by a 20% or more change in main exposure odds ratios when potential confounders were included in the model.

**Fig 1:**
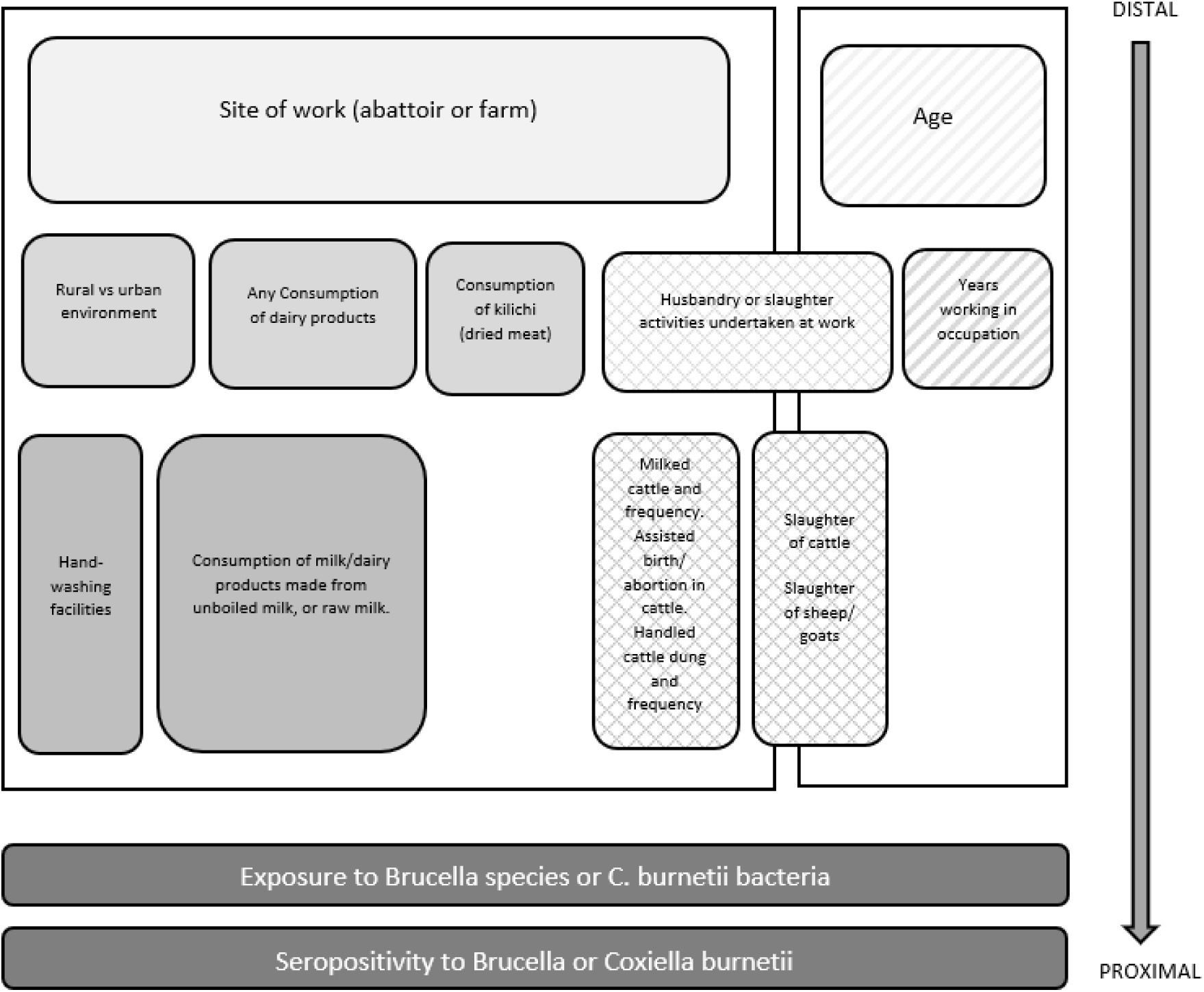
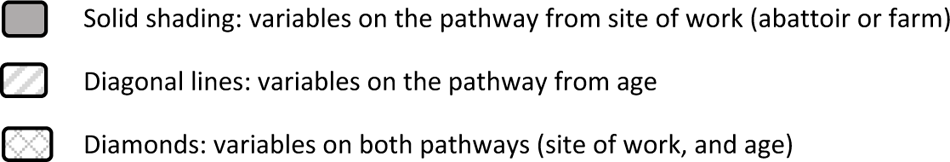
Hierarchical conceptual framework of the risk factors for seropositivity for *Brucella* and *Coxiella burnettii*. Legend: Shading becomes darker the more proximal a variable is to the outcome.

## Results

### Participant characteristics

Overall, 189 participants were recruited including 108 abattoir workers, and 81 farmworkers from 52 dairy farms. Three abattoir workers refused to take part. Farm workers tended to be younger than the abattoir workers with less formal education (Table1). While eight abattoir workers also did some work on farms, no farmworker worked in abattoirs.

### Dairy farm characteristics

There were high-levels of livestock ill health on farms, including ruminant abortions and cattle hygromas (Table 2). Fifty-one (98.1%) farms gave bulk milk samples, of which 20 (39.2%, 95% CI 26.6 - 53.4%) tested positive for Brucella.

**Table 1:**
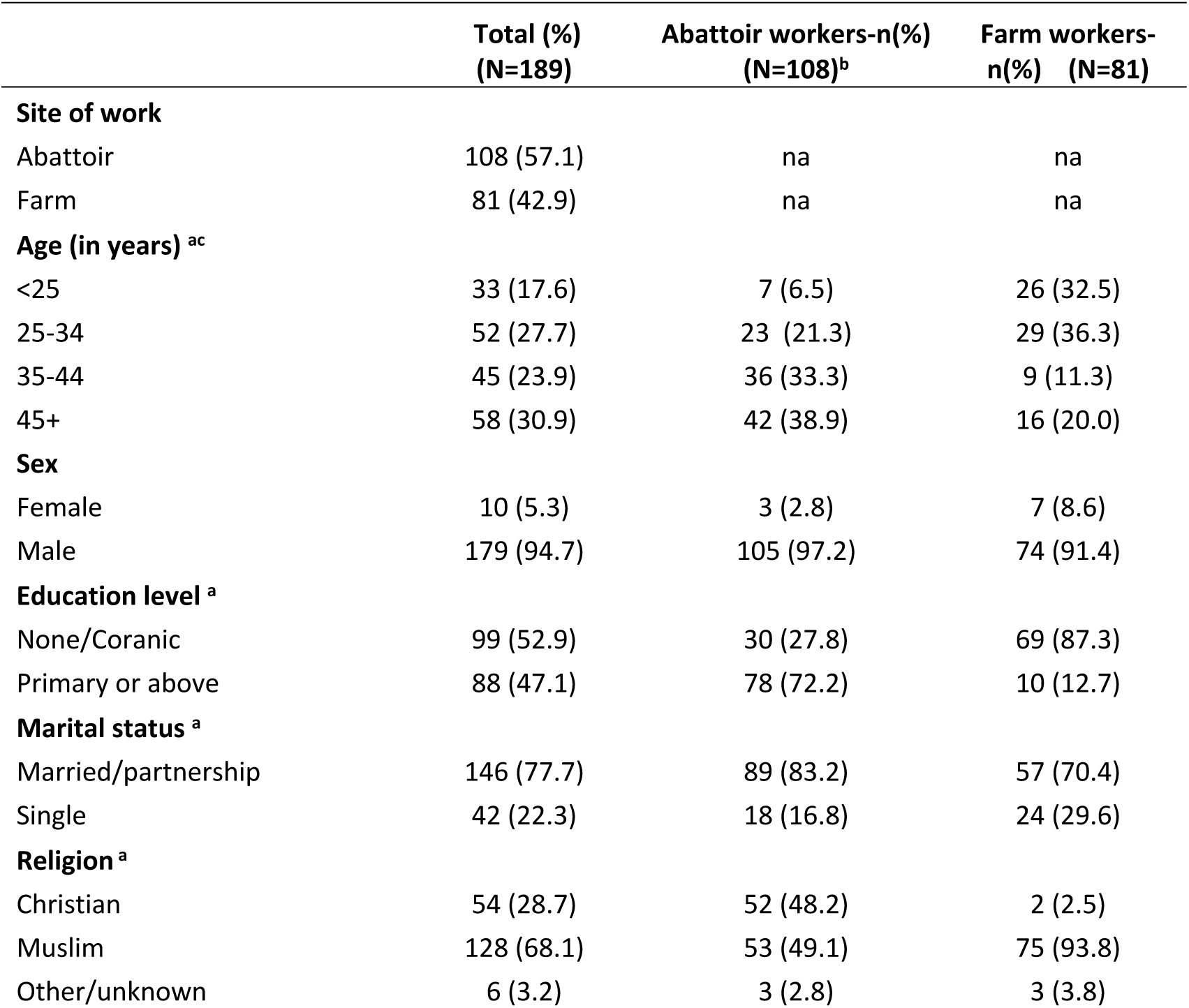
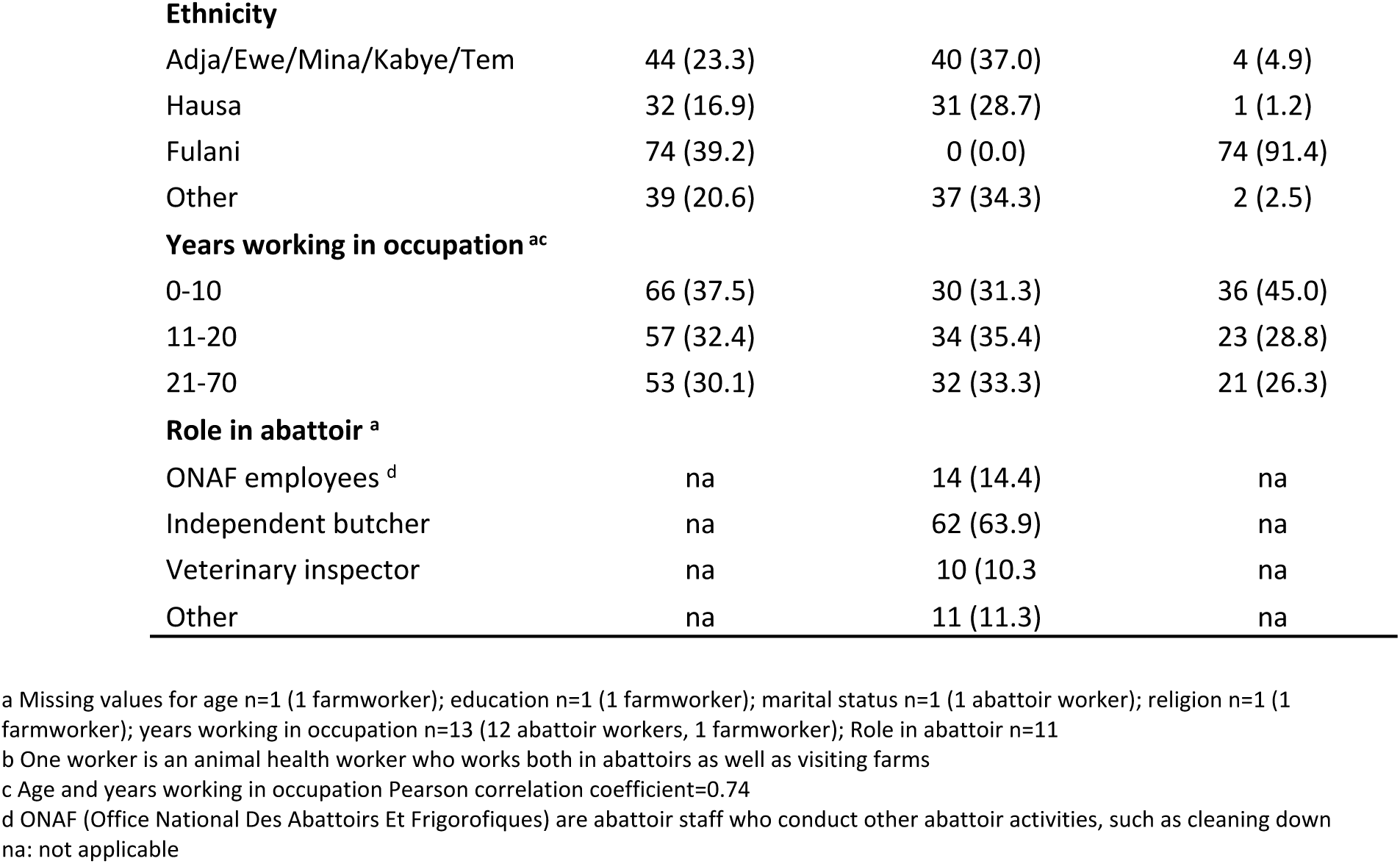
Characteristics of the study population, both overall and by site of work.

**Table 2:**
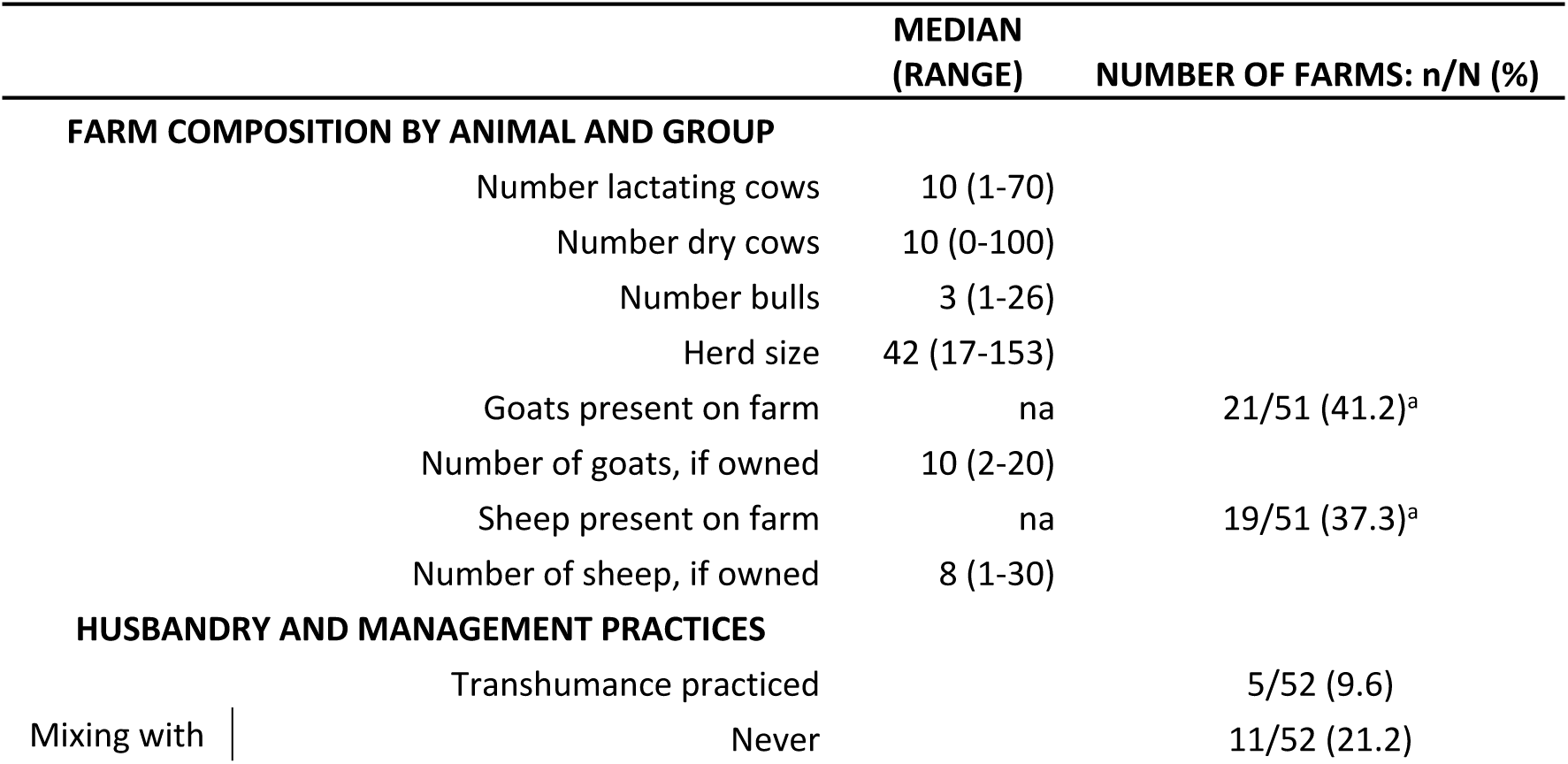
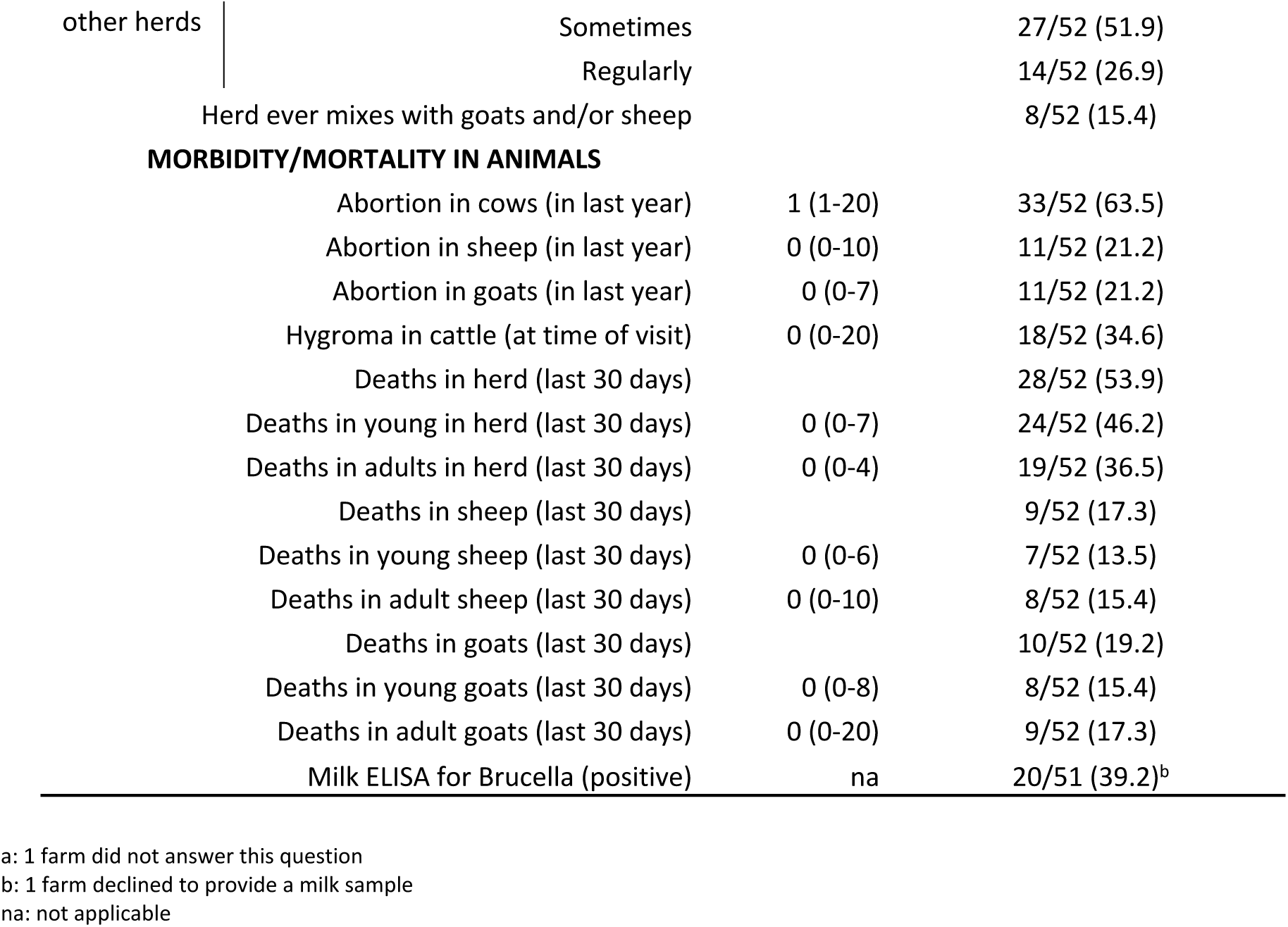
Farm characteristics of the 52 participating dairy farms: a) farm composition by animal and group, b) husbandry and management practices, c) morbidity and mortality in animals.

### Seroprevalence against *Coxiella burnetii* and *Brucella* species (S1 Table)

Seven abattoir workers (6.5% (95% CI: 5.1-8.2)) and 11 farmworkers (13.6 % (95% CI: 6.9-24.9)), were seropositive for *Brucella*. *Coxiella* seroprevalence was higher at 31.5% (95% CI 24.6-39.3) in abattoir workers and 23.5% (95% CI: 15.4-34.1) in farmworkers. Eight (4.2%) participants were positive for both pathogens, including six farmworkers.

### Overall animal contacts at the individual level (S2 Table)

Forty-one percent of abattoir workers and 95% of farm workers took part in animal husbandry in the last year. The majority of both abattoir and farm workers had been involved in cattle slaughter while more than 30% had slaughtered small ruminants in the past. Of the 79 abattoir workers who took part in slaughter only 5% wore protective equipment to protect their eyes, hands or mouth during husbandry and slaughter activities with ruminants and only 4% of farmworkers did so during husbandry activities with ruminants.

### Consumption of dairy and other livestock products (S3 Table)

More than two-thirds of participants consumed cheese (78.3%), milk (76.7%), yoghurt (68.3%), and fermented milk (66.7%). More people ate dairy products made from boiled cow’s milk than from unboiled at least once a month (84/189 (44%) and 56/189 (30%) respectively). The majority reported consumption of unboiled (57%) and boiled milk (56%) on at least a monthly basis. History of consumption of milk (12%) or dairy products (16%) from small ruminants was not common.

### Risk factors for Brucella seropositivity

No variable was associated with Brucella seropositivity in the abattoir group (Table 3).

**Table 3:**
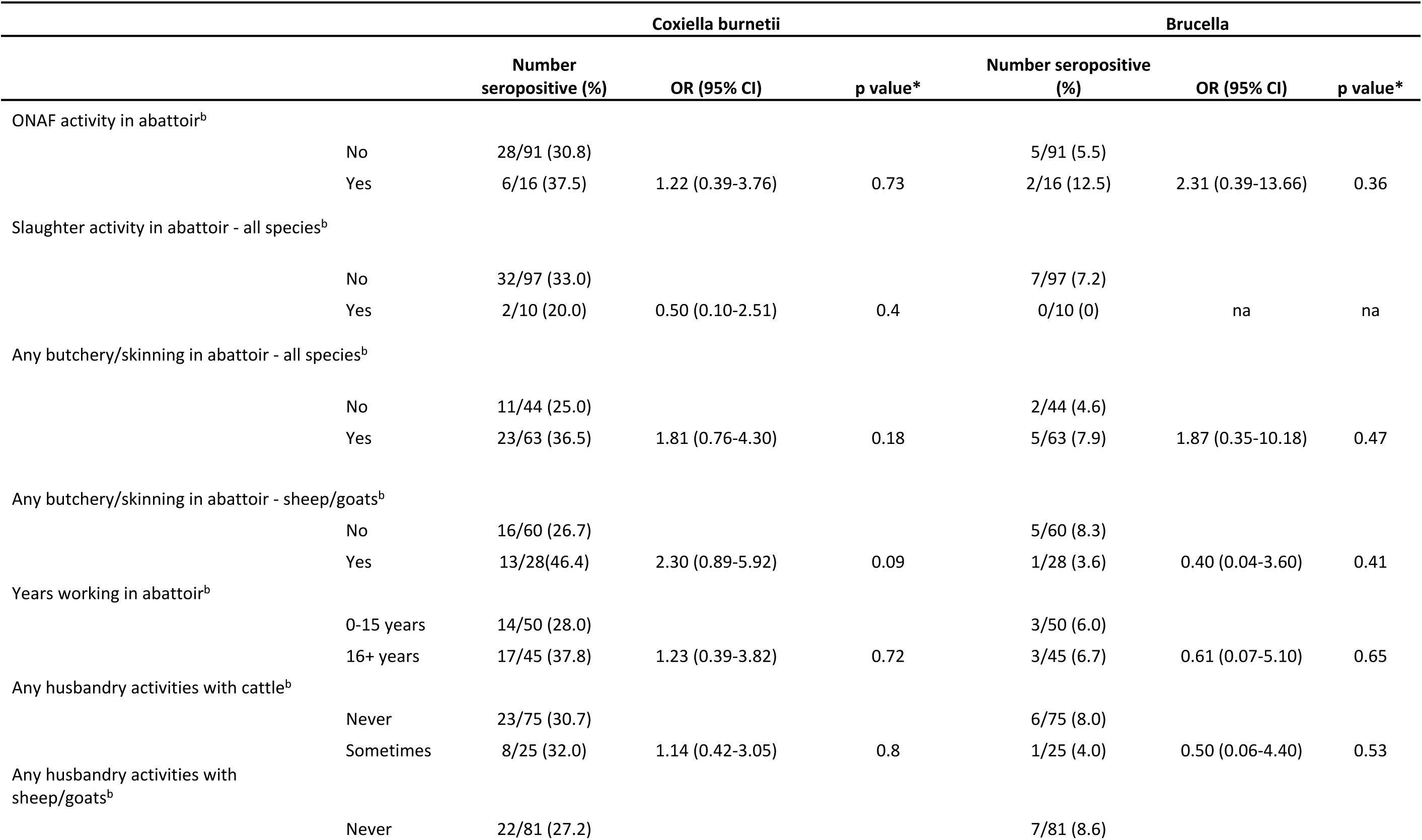
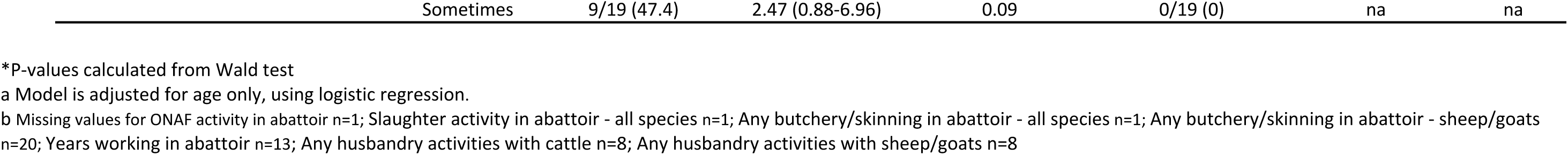
Age-adjusted associations^a^ between animal-contact risk factors and Brucella and Coxiella seropositivity in abattoir workers.

There was some evidence that animal ill health on farms was associated with Brucella seropositivity in farmworkers (abortion in goats OR 5.22, 95% CI: 1.01-26.99, p=0.05; death of young cattle OR 6.42 95% CI: 0.72-57.40, p=0.1; death of young goats OR 5.81 95%CI: 1.0-33.81, p=0.05) (Table 4). The odds of human seropositivity were five times higher on farms with a positive milk test than those with a negative test (OR 5.15, 95% CI: 1.21-21.97, p=0.03).

**Table 4:**
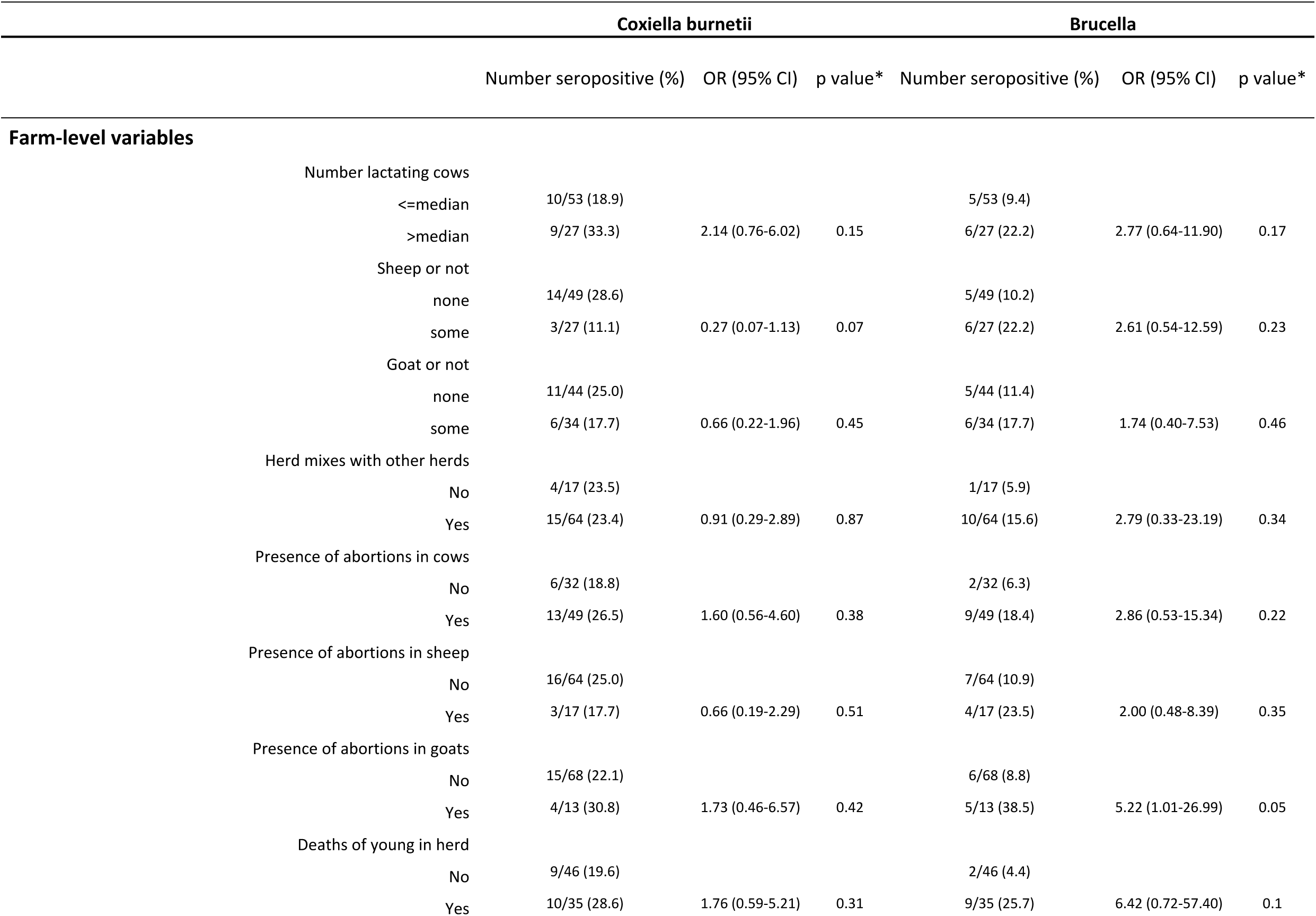
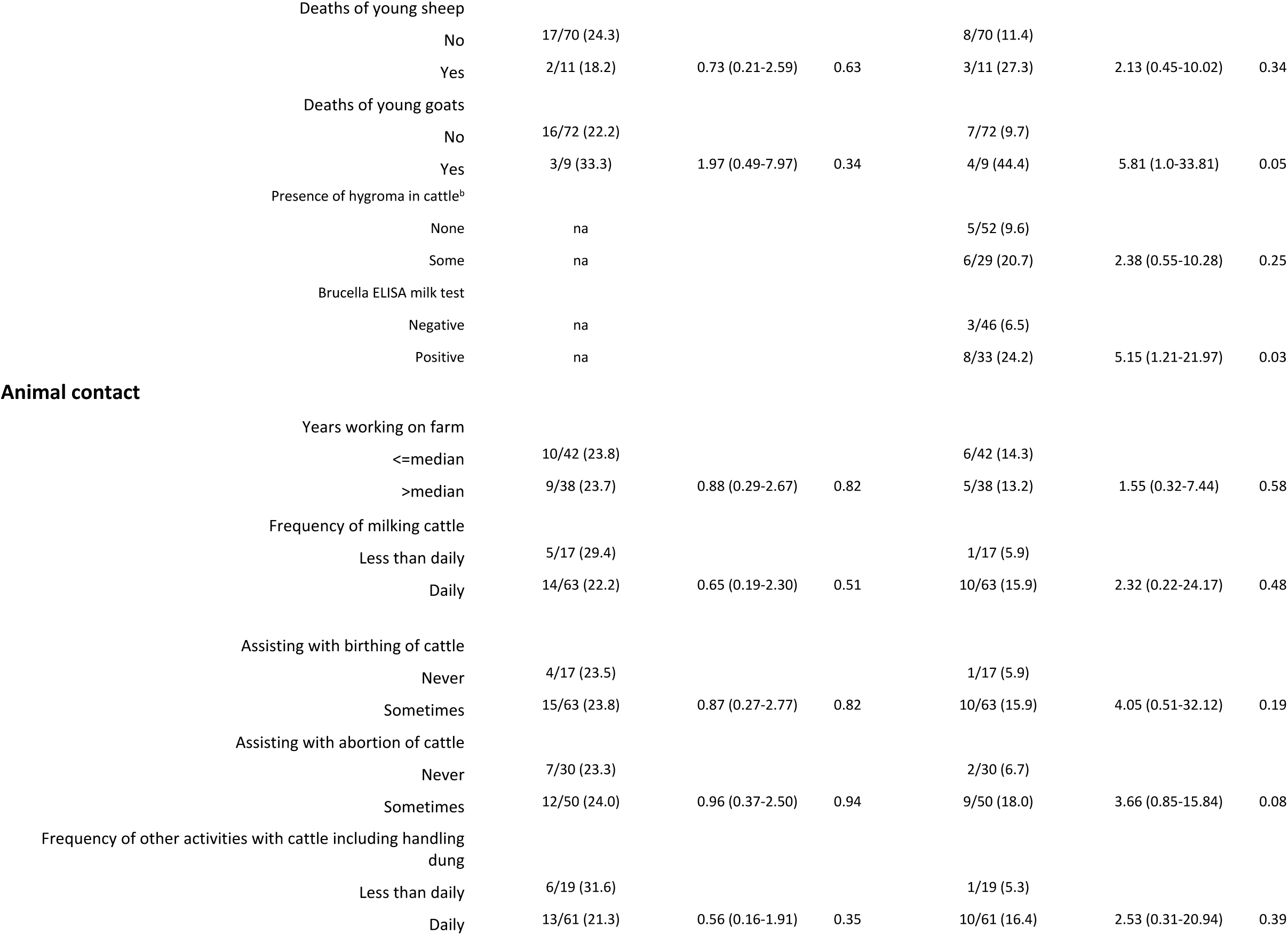
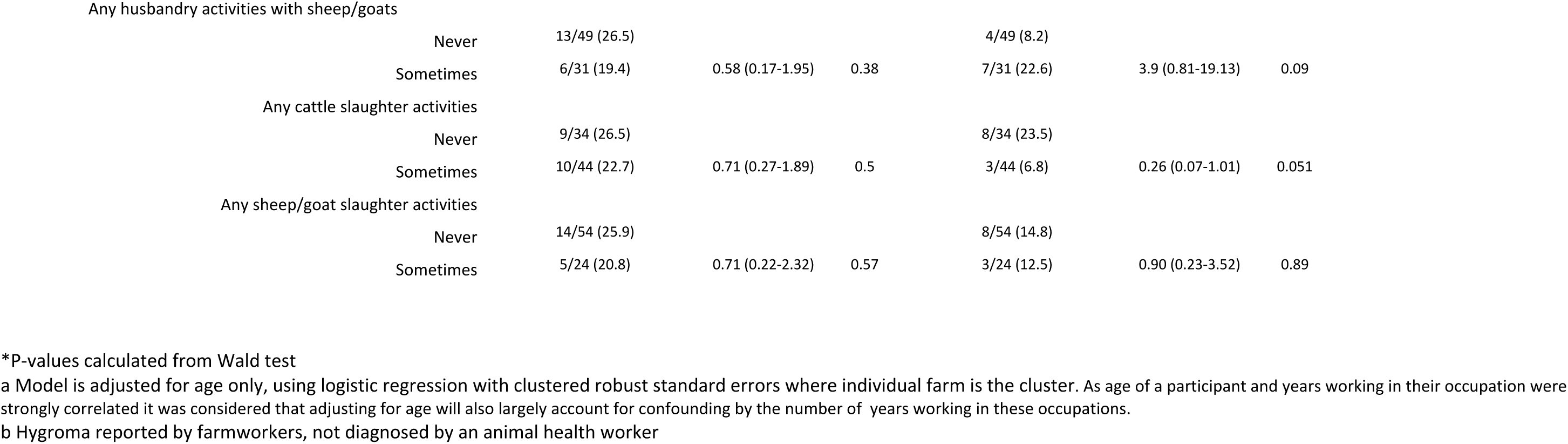
Age-adjusted associations^a^ between farm-level and animal husbandry risk factors and Brucella and Coxiella seropositivity in farmworkers.

There was weak evidence that participating in small ruminant husbandry was associated with nearly four times higher odds of Brucella seropositivity (OR 3.9 95% CI 0.81-19.13, p=0.09), as did assisting with cattle abortions (OR 3.66, 95%CI: 0.85-15.84, p=0.08).

Farmworkers had nearly twice the odds of being seropositive compared to abattoir workers (OR 1.93, 95% CI: 0.94-3.93, p=0.07). Consuming unboiled milk on a monthly basis or more was associated with higher odds of Brucella seropositivity (OR 3.79, 95% CI: 2.34-6.13, p<0.001) (Table 5).

**Table 5:** Associations between socio-demographic and dairy consumption factors and seropositivity to the two pathogens Brucella species and Coxiella burnetii in all participants^a^.

|  |  | *Coxiella burnetii* |  |  | *Brucella* |  |  |
| --- | --- | --- | --- | --- | --- | --- | --- |
|  |  | Number positive (%) | OR (95% CI) | p value* | Number positive (%) | OR (95% CI) | p value* |
| **Education level** | None/coranic | 25/99 (25.3) |  |  | 14/99 (14.1) |  |  |
|  | Primary or above | 28/89 (31.5) | 1.11 (0.64-1.94) | 0.71 | 4/89 (4.5) | 0.34 (0.19-0.61) | <0.001 |
| **Site type** | Abattoir | 34/108 (31.5) |  |  | 7/108 (6.5) |  |  |
|  | Farm | 19/81 (23.5) | 0.76 (0.45-1.29) | 0.32 | 11/81 (13.6) | 1.93 (0.94-3.93) | 0.07 |
| **Consume fermented milk including Caille** | no | 16/63 (25.4) |  |  | 4/63 (6.4) |  |  |
|  | yes | 37/126 (29.4) | 1.64 (1.22-2.19) | 0.001 | 14/126 (11.1) | 1.29 (0.58-2.90) | 0.53 |
| **Consume cheese** | no | 9/41 (22.0) |  |  | 3/41 (7.3) |  |  |
|  | yes | 44/148 (29.7) | 1.69 (0.98-2.92) | 0.06 | 15/148 (10.1) | 1.27 (0.47-3.38) | 0.64 |
| **Consume yoghurt** | no | 15/60 (25.0) |  |  | 5/60 (8.3) |  |  |
|  | yes | 38/129 (29.5) | 1.30 (0.83-2.04) | 0.25 | 13/129 (10.1) | 1.32 (0.52-3.36) | 0.56 |
| **Consume unboiled cows dairy products** | Less than monthly | 38/133 (28.6) |  |  | 12/133 (9.0) |  |  |
|  | At least monthly | 15/56 (26.8) | 1.09 (0.47-2.53) | 0.83 | 6/56 (10.7) | 0.91 (0.29-2.88) | 0.88 |
| **Consume unboiled cows milk** | Less than monthly | 24/81 (29.6) |  |  | 3/81 (3.7) |  |  |
|  | At least monthly | 29/108 (26.9) | 1.12 (0.8-1.58) | 0.5 | 15/108 (13.9) | 3.79 (2.34-6.13) | <0.001 |
| **Frequency of consuming any cows milk** | Less than monthly | 17/54 (31.5) |  |  | 2/54 (3.7) |  |  |
|  | Monthly | 36/135 (26.7) | 0.97 (0.74-1.28) | 0.74 | 16/135 (11.9) | 2.63 (1.97-3.49) | <0.001 |
| **Consume kilichi** | Less than monthly | 26/98 (26.5) |  |  | 12/98 (12.2) |  |  |
|  | At least monthly | 26/90 (28.9) | 1.03 (0.66-1.61) | 0.89 | 6/90 (6.7) | 0.57 (0.22-1.46) | 0.24 |
| **Consume any milk/dairy from sheep/goats** | No | 27/88 (30.7) |  |  | 6/88 (6.8) |  |  |
|  | Yes | 26/101 (25.7) | 0.76 (0.52-1.11) | 0.16 | 12/101 (11.9) | 0.48 (0.22-1.05) | 0.07 |

\* P-values calculated from Wald test
a Model is adjusted for age and type of site of work (abattoir or farm), using logistic regression with clustered robust standard errors where site of work is the cluster. No interaction was observed between the site type of participants and any of the risk factors examined

### Risk factors for Coxiella seropositivity

Adjusting for age and considering farm-level clustering, there was no evidence of an association with any livestock husbandry or livestock health variable in farmworkers (Table 4).

After adjusting for age only, there was no evidence that abattoir activities were associated with seropositivity, other than weak evidence of an association with butchery/skinning of small ruminants (p=0.09) or in participating in any husbandry with sheep/goats (OR 2.47, 95% CI 0.88-6.96, p=0.09) (Table 3).

The odds of Coxiella seropositivity in those who consumed fermented milk (OR 1.59, 95% CI: 1.26-2.00, p<0.001) and in those who consumed cheese (OR 1.70, 95% CI: 0.97-2.98, p=0.07) were greater than in those who did not when looking at all participants in a multivariable model considering dairy product consumption (Table 6).

**Table 6:** Multivariable^a^ model for consumption variables associated with Coxiella seropositivity in all participants.

| Variable | Adjusted OR (95% CI) | p-value* |
| --- | --- | --- |
| **Consume fermented milk including Caille** |  |  |
| No | 1 |  |
| Yes | 1.59 (1.26-2.00) | <0.001 |
| **Consume cheese** |  |  |
| No | 1 |  |
| Yes | 1.70 (0.97-2.98) | 0.066 |
| **Frequency of consuming any cow's milk** |  |  |
| Less than monthly/never | 1 |  |
| Monthly | 0.73 (0.54-0.98) | 0.002 |
| **Ever consumed sheep/goat's milk or dairy products** |  |  |
| No | 1 |  |
| Yes | 0.75 (0.53-1.06) | 0.099 |
| **Type of site** |  |  |
| Abattoir | 1 |  |
| Farm | 0.66 (0.37-1.19) | 0.17 |

\*P-values calculated from Wald test a Logistic regression model includes variables Consumption of fermented milk including Caille, Consumption of cheese, Frequency of consuming any cow's milk, Ever consumed sheep/goat's milk or dairy products, and the *a priori* variables age and type of site and clustered robust standard errors where site of work is the cluster No interaction between exposure variables and whether a worker's site of work was an abattoir or farm was seen.

### Fever history and related behaviours in all workers

Fever in the last year was commonly reported in the livestock workers (149/188, 79.3%) (S4 Table). Participants reporting fever in the last month were three times more likely to be Brucella seropositive (OR 3.63, 95% CI 1.21-10.90, p=0.014) and no fever-related variable was associated with Coxiella seropositivity. The majority of participants who had experienced fever in the last year received no testing (82/141) or only for malaria (59/141). Antimalarials (76/142) and antibiotics (55/142) were commonly taken as treatment. Those seropositive for Brucella were highly likely to have taken antimalarials during a fever in the last year (68.8%) compared to those who were seronegative (51.6%) (p=0.044).

## Discussion

We found, through a cross-sectional study of livestock workers, that *Brucella* and *Coxiella burnetii* were present concurrently in people with occupational exposure in peri-urban Lomé with overall human seroprevalences of 9.5% and 28% respectively. Seroprevalence varied with site type, with a higher seroprevalence to *Brucella* on farms (13.6% compared to 6.5% in abattoirs) and to *C.burnetii* in abattoir workers (31.5% compared to 23.5% on farms). Bulk milk samples from associated cattle herds found that 39% (95% CI 27-53%) of farms also had Brucella-positive milk samples, lower than a previous study in Lomé’s peri-urban herds in 2017(62%, 95%CI 55-69%) [12]. For both diseases there was an association with the consumption of dairy products (unboiled milk in the case of Brucella and fermented milk and cheese in the case of Q fever), and ruminant husbandry (small ruminant husbandry and assisting with cattle abortion in farmworkers for Brucella, and small ruminant husbandry in abattoir workers for Q fever). The odds of Brucellosis seropositivity in farmworkers was also increased with morbidity/mortality of owned ruminants (death of young cattle and goats and abortion in goats). Considering this, the widespread ill-health in livestock including death of young animals and abortions which are consistent with Brucellosis/Coxiellosis, consumption of unboiled milk and dairy products, and lack of PPE usage is of concern.

Heterogenous incidence of both diseases in human and animal populations, both between and within low and middle income countries, and over time, means that it is difficult to extrapolate from previous studies [7,14]. Our findings on Brucella seroprevalence were in contrast to a 2013 study in Northern Togo which found a seroprevalence of 2.4% in Fulani villagers, a comparable population to the farm workers in this study who had a seroprevalence of 13.6% [13]. This may be due to a lower prevalence in animals (9.2% in village cattle, 7.3% in transhumant cattle and 0% in small ruminants) though these results are from individual serology rather than bulk milk sampling [13]. The higher seroprevalence to *Coxiella burnetii* we note is aligned with the same study which found more than ten times the prevalence against *Coxiella* compared to *Brucella* in both Fulani and non-Fulani groups [13]. Similar, though not as marked, findings were seen in studies in Ethiopia, and Kenya [15,16,18]. Despite a higher seroprevalence of *Coxiella burnetii* in many African contexts, Brucellosis is often given greater attention by health agencies and research funders [19,20]. Q fever can have severe health sequelae, being responsible for up to 5% of endocarditis cases globally, and if this disparity in seroprevalence is reflected in clinical burden then it is important to reassess Q fever as a priority [21].

Q fever had a seroprevalence of 31.5% in the abattoir workers, despite the lack of abattoir-specific activities being associated with seropositivity. This was in line with de Boni et al who found no specific abattoir activity was a risk factor and hypothesised that workers were exposed when the bacterium was aerosolized during slaughterhouse activities [22]. Aerosols are a major transmission mode for *Coxiella burnetii* and this abattoir with its lack of ventilation provides ideal conditions for exposure of employees regardless of role [9]. The finding that Dutch cull workers were at greater risk of seroconverting if they worked indoors rather than outdoors supports this [23].

For both diseases infected livestock are the main reservoir, shedding bacteria in birth and abortion materials, milk, faeces, and urine [8,9]. Close contact with ruminants has been shown to be associated with seropositivity in Brucellosis and Q fever, as was seen in our study in those who participated in small ruminant husbandry and assisting with cattle abortions [24,25].

Abortion, birth of weak offspring and death of neonatal animals are symptoms of ruminant Brucellosis and Coxiellosis [26,27]. We demonstrated associations between human Brucella seropositivity and livestock ill-health markers (abortion in goats, and the death of young cattle on farm). Literature on this is limited and largely confined to animal abortion [28]. Animal-health variables such as abortion, and mortality in neonatal animals, may be indicators of herd/flock infection, and could be monitored using syndromic surveillance to mitigate risk to workers and livelihood losses. The association between Brucella seropositivity with presence of abortion in goats, and with participating in small ruminant husbandry is of particular interest as we know *B.melitensis* is circulating in ruminants in this region (personal communication). This highlights the complexity of transmission amongst species and to humans and the need for multi-species studies.

This population consumed a high-level of bovine dairy products, including those sourced from unboiled milk. We found both consuming fermented milk and cheese were associated with *Coxiella burnetii* seropositivity. A Latvian study published in 2021 found a high prevalence of *C.burnetii* DNA in these products, showing they are viable transmission vehicles [29]. Previously dairy products were considered to rarely contribute to *C.burnetii* transmission but the evidence is mixed with some studies finding an association [2,13,30].

Regular unboiled milk consumption increased the odds of Brucella seropositivity. Multiple studies have found consumption of unpasteurised milk to be a risk factor [31–33]. Consumption of small ruminant dairy products has also been shown to be a risk factor for Brucellosis [34,35]. However, consumption of small ruminant dairy products isn’t culturally common in Togo, despite large numbers of these species, and those who did consume these weren’t found to have increased odds of seropositivity.

Sampling frames for occupational groups in LICs are unlikely to pre-exist and without a comprehensive census sampling a representative group is a challenge. There are also practical limitations in carrying out a questionnaire-based survey in occupational groups with limited time for participation. Dean et al regarded a lack of detailed information on exposures as a limitation in thoroughly assessing risk [13]. While our questionnaire allowed more nuanced examination of risk factors it also limited the sample size. For this reason we could not perform certain analyses such as adjusting for other exposures of interest, including other animal husbandry and dairy consumption variables, in multivariable analysis for Brucella seropositivity.

There is a paucity of studies looking at linked animal and human data. Here we were able to link human data on Brucellosis to milk sourced from associated cattle to show an association between presence of a positive bulk milk sample and human seropositivity. Bulk milk sampling is relatively affordable and easy and could be utilised in regular herd surveillance. To expand on this study, future studies which conduct direct multi-species sampling including small ruminants with, in the case of *Brucella*, speciation of any isolated bacteria would improve understanding of how the organisms are cycling through the ecosystem.

There were low levels of PPE use in both groups. Many studies, though not all, have found PPE to be beneficial in protecting against both diseases [36,37]. PPE use should be recommended, particularly adequate masks (P2/N95) in abattoir areas where aerosols are produced [38].

This population is affected by both zoonoses, and there is a need for preventative measures. Implementation should involve at-risk workers, trusted community leaders, health practitioners and policymakers using culturally sensitive, pragmatic and economically feasible approaches. Our study highlights the need for increased clinician awareness about zoonoses, and for relevant history taking including occupation, which might raise clinical suspicion, particularly as malaria prevalence declines [39]. Seropositive participants commonly experienced febrile disease though no symptoms were found to be associated, and this non-specific clinical picture hinders diagnosis by clinicians. Many seropositive participants were only tested for malaria and treated with antimalarials and non-specific antibiotics, similar to other studies [40, 41]. In order to improve diagnosis healthworkers require resources to test and treat non-malarial illnesses, both to improve clinical outcomes and to reduce inappropriate prescribing which contributes to selection for antimicrobial resistance.

The need for complex interventions involving interdisciplinary public health teams is exacerbated in zoonoses [19]. However, integrated control of multiple zoonotic diseases, such as Q fever and Brucellosis, through interventions targeting both the animal and human populations could have a dual benefit through improving animal health and livelihoods, while simultaneously decreasing human disease, to alleviate the cycle of poverty. Such integrated approaches would be more cost-effective by optimising resource use, especially when working with marginalised communities who are isolated from public services [42]. The findings of our study provide evidence which can be used to tailor such public health interventions against both Brucella and Q fever in occupational groups working with livestock.

## Data Availability

The data that supports the findings of this study will be publicly available from the London School of Hygiene and Tropical Medicine public data repository accessible at https://datacompass.lshtm.ac.uk/.

## Acknowledgements

We thank all the livestock workers and abattoir workers from the municipal abattoir in Lomé who participated in this study, and community leaders who facilitated this study. This study was part of a larger UKRI Zoonosis in Emerging Livestock Systems project and we thank the PI of this study Javier Guitian, as well as Ayayi Akakpo and Imadidden Musallam for facilitating the study. We thank Wemboo Afiwavi Halatoko, Koffi Akolly, Zoulkarneiri Issa, Adjaho Komla Koba and Afeignidou Telou at the INH and Emilie Go-Maro, Rufine Yempab, and Komlan Mawufemo Adjabli at the Department de l’Elevage who were involved in processing, testing and storage of samples. Our appreciation goes to the zootechnicians of the Department de l’Elevage Koffivi Amewou, Kom Agbéssi Adjessoklou and Dossou Zanou who facilitated all visits to dairy farms. Thanks also go to the field team; Kodjo Afangbegnon, Robertine Amoussou and Kocou Jean-Paul Amoussou.

## Supporting Information captions

**S1 Table:** Serological results of each test for *Brucella* species and *Coxiella burnetii*

**S2 Table:** Livestock contact, including by site of work

**S3 Table:** Dairy product and kilichi consumption, including by site of work

**S4 Table:** Health and health seeking behaviours in participants in the previous 12 months

